# Wearable sensor data characterizes vigilance and avoidance behaviors in young children with mental health symptoms during a threat induction task

**DOI:** 10.64898/2026.03.31.26349900

**Authors:** Jenna G. Cohen, Guido Mascia, Bryn C. Loftness, Mary Carter Bradshaw, Julia Halvorson-Phelan, Josh Cherian, Dheeraj Kairamkonda, David C. Jangraw, Ryan S. McGinnis, Ellen W. McGinnis

**Author notes:** This research was funded by the US National Institutes of Health (MH123031) and the US National Science Foundation (2422226).

## Abstract

Early childhood mental health problems are common and difficult to detect due to reliance on caregiver reports of often unobservable symptoms. This study quantified threat response movement patterns during a 30-second laboratory threat induction task using wearable inertial sensors. Movement patterns were used to examine (1) changes in stimuli response across the task (task validity) and (2) associations with symptom severity (clinical validity). Sacral accelerometer and gyroscope data were analyzed from 91 children aged 4-8 years during the brief task, 48.4% of whom had a mental health diagnosis. Consistent with task validity, Turning Speed varied across task phases differing in potential threat intensity. Consistent with clinical validity, internalizing symptoms were associated with smaller Turning Angle, possibly indicating vigilance. This effect was moderated by comorbid externalizing symptoms, such that children with high internalizing and high externalizing symptoms exhibited larger Turning Angles, possibly indicating avoidance. Findings demonstrate that brief wearable-enabled tasks can capture subtle objective behavioral markers of threat responses and underscore the importance of considering comorbid symptom dimensions in early childhood mental health screening.

## Introduction

Childhood mental health problems are prevalent and often emerge early in life (1,2). More than one in five children in the United States have a mental health diagnosis (1,2), and children under six years of age experience the highest rates of unmet mental health needs (3). Early detection is critical, as untreated emotional and behavioral problems during childhood increase the risk of adverse health outcomes later in life (4–6).

Since young children are often unreliable reporters of their own internal states (7–9), recognizing and reporting symptoms of mental health problems typically falls to their caregivers. Diagnoses are often based on caregiver report surveys (i.e., (10)) and semi-structured clinical interviews (i.e., (11)). However, even attentive and well-intentioned caregivers may under-(12) or over-report (13–15) symptoms, as many symptoms are not directly observable (e.g., feeling sad or fearful (3)), and caregivers’ own mental health experiences may influence their perceptions of their children’s behaviors (13–15). As a result, there is a critical need for objective tools that can complement caregiver reports in screening for childhood mental health risk.

In research settings, laboratory-based stress paradigms have been used to examine children’s behavioral and physiological responses to emotionally stimulating or threatening events (16–21). Paradigms inducing anxiety or fear for young children are often behaviorally coded for negative affect (22) and behavioral inhibition (23), or in the case of adults, fear-potentiated startle response (24), with each showing associations with current or subsequent anxiety and depressive disorders. Theories posit that individuals with anxiety and depressive disorders exhibit cognitive biases (20,25–27) in how they attend and interpret information through both automatic and strategic processes (21,26,28,29). Evidence for these theories has typically been measured using eye tracking metrics during computerized threat paradigms (21,30) and demonstrates that children with anxiety disorders and/or high anxiety symptoms orient toward external threatening stimuli more than children without (21,26,31), which is interpreted as vigilance.

In comparison to anxiety, other mental health conditions exhibit distinct, often opposing threat responses. For instance, Attention Deficit Hyperactive Disorder (ADHD) has been associated with a blunted response when anxiety is associated with heightened response measured via stress hormones (cortisol (32,33)), as well as slower orientation to threatening faces when anxiety is linked with faster orientation. However, despite opposing threat responses across internalizing and externalizing problems (e.g., (19,34–38)), and prevalent (roughly 25-50% of cases (1)) heterotypic (internalizing and externalizing) comorbidity in children (34), direct comparisons for children within studies are rare. One such study (n=27 children) confirmed that ADHD and anxiety symptoms exhibited opposing threat responses, but interestingly, showed that in children with high co-occurring symptoms, the vigilant anxiety response prevailed (19). Studies like this must be replicated, as focusing on anxiety and depression (internalizing symptoms) without considering ADHD and/or oppositional defiance (externalizing symptoms) limits the ability to generalize results to real-world pediatric populations.

The Research Domain Criteria (RDoC) (39) provides a framework for examining the underlying biology and behaviors of mental health using dimensional systems that better capture variability in symptom expression (40,41). For instance, measuring how individuals process and respond to Potential Threat, one construct within the Negative Valence System, or how humans process negative emotions may aid understanding of their functional impairment (24,39). A paradigm used to measure response to Potential Threat is the Approach task, wherein child movement (orienting toward or away from a stimulus) can be measured during 30 seconds while they are led into a darkened room to an unknown object (a box covered with a blanket) (24). This brief behavioral task using one wearable sensor (with an accelerometer and gyroscope) could increase feasibility compared to eye tracking paradigms which require sustained child attention, as well as specialized lab equipment and software.

In the current study, we use a wearable inertial sensor to characterize movement during the Approach Task in children across a range of internalizing and externalizing symptom severity. We aim to use child movement measurements to examine (1) changes in threat response across the task (task validity) and (2) associations with symptom severity (clinical validity). We hypothesize that (1) child movement will differ over time across the task, reflecting expected threat-related responses according to intensity and modulation, that (2) child movement will be associated with internalizing symptom severity, and that (3) this association will be moderated by comorbid externalizing symptom severity.

## Methods

### Participants and experimental protocol

One hundred and four children (female 38.5%, White non-Hispanic 89.4%) aged 4-8 years (mean ± standard deviation: 81.4 ± 14.5 months) with (47.6%) and without (52.4%) mental health diagnosis were recruited via digital community advertisements (n=98) and the Vermont Center for Children, Youth and Families (n=6) for a one-time, 2.5-hour laboratory visit. The University of Vermont Institutional Review Board approved this study (CHRBSS 00001218), and all participating families gave their signed consent. Exclusion criteria consisted of having a suspected or diagnosed developmental disorder or serious medical condition (e.g., diabetes, cardiovascular disease). This work is part of a larger study of potential biomarkers of childhood mental health (17,37).

At the start of the laboratory visit, after obtaining consent, the child was equipped with wearable sensors, including a BioStamp nPoint® sensor secured with adhesive directly to the skin of the lower back at approximately the L5 level (Figure 1). We refer to this as a sacral device placement and consider data from this location in this work as it provides a suitable summary of whole-body movements. The child was then accompanied by a research assistant to a separate room to complete three mood induction tasks, each separated by 2-to 3-minute rest periods. The tasks included (1) a 30-second Approach Task in which the child followed the research assistant into a dimly lit room, (2) a 3-minute modified version of the Trier Social Stress Test for Children (TSST-C) where the child was instructed to improvise a story in front of an emotionless judge, and a 3-minute Bubbles Task during which the child was encouraged to play with bubbles (17,42).

**Fig 1.**
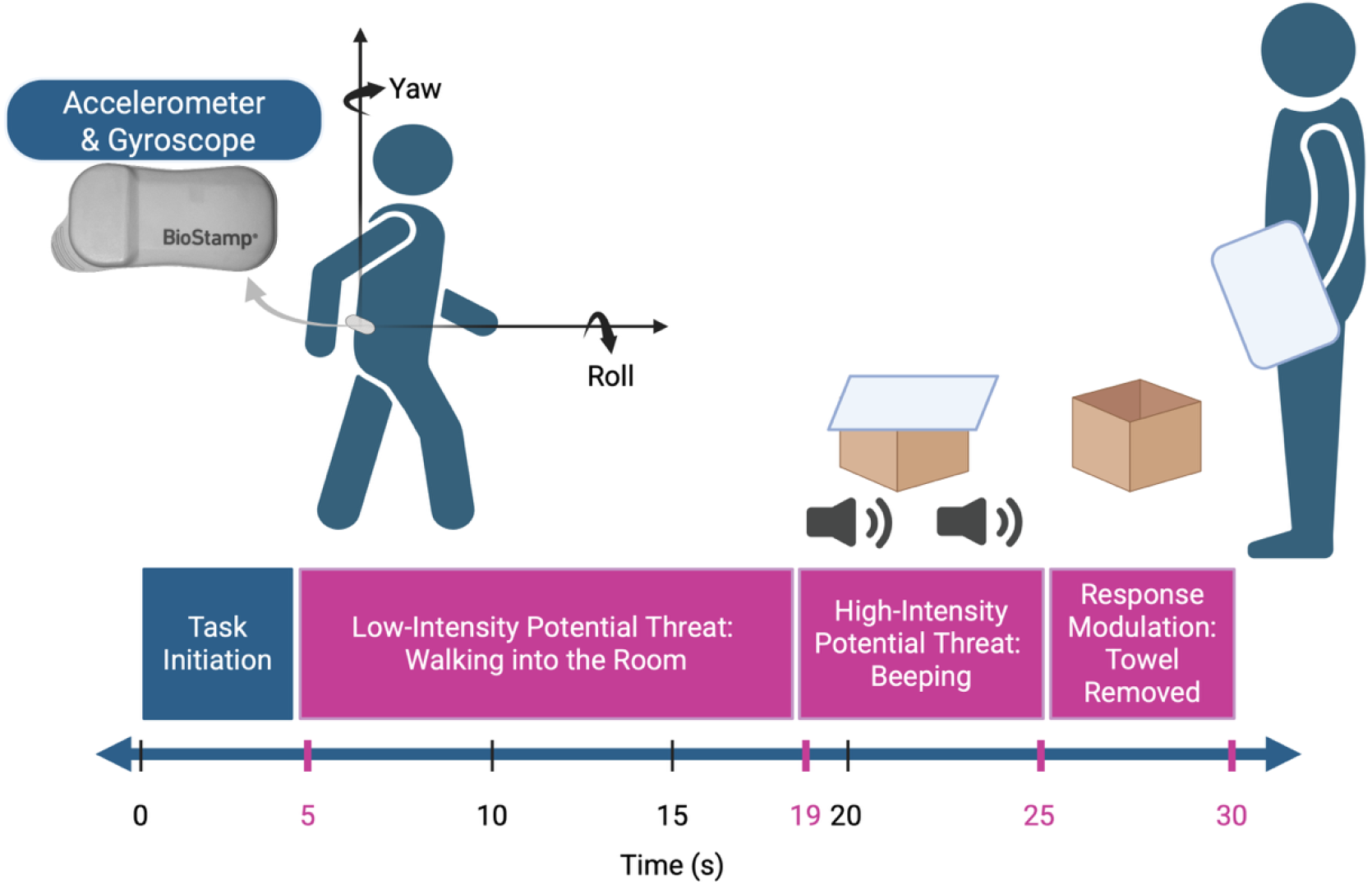
Thirty-second stress task. Sacrum orientation features were considered during the full task and the following three phases: Low-Intensity Potential Threat (5-19 seconds), High-Intensity Potential Threat (19-25 seconds), and Response Modulation (25-30 seconds).

While the child completed the tasks, caregivers completed surveys including the Child Behavior Checklist (CBCL) (10) to quantify children’s emotional and behavioral problems. Internalizing and externalizing T-scores of the CBCL were derived according to standard scoring procedures and used as continuous indices of symptom severity for this analysis. This dimensional approach aligns with the RDoC framework, which emphasizes continuous, mechanism-oriented measures rather than categorical diagnoses, particularly for young children (7–9).

Of the 104 child participants, two children refused to wear BioStamp sensors, five children did not have the necessary timestamp data as collected from a smartphone due to technical issues, and six children did not have data from the sacrum sensor during the baseline period needed for calibration. Thus, we consider data from 91 children for analysis. Child demographics did not differ by wearable sensor data availability (age: *χ*^*2*^ = 46.10, *df* = 46, *p* = .468; white non-Hispanic vs other: *χ*^*2*^ = 0.13, *df* = 1, *p* = .718; male vs female: *χ*^*2*^ = 0.37, *df* = 1, *p* = .542).

### Approach Task for young children

The Approach Task (adapted from (36)) is a 30-second task, wherein the child was led by an administrator into an unfamiliar, darkened clinic room toward an unknown object (which is actually just an empty box) hidden underneath a blanket (Fig 1). The child begins the task facing the closed door into the clinic room. From 0 to 5 seconds into the task the administrator opens the clinic room door and ushers the child inside, stating “I have something to show you,” and is defined as the Task Initiation phase. From 5 to 19 seconds, the child was led into the room slowly toward an ambiguous object. This phase is meant to induce Low-Intensity Potential Threat responses such as vigilance and anticipatory anxiety (36,41), and is defined as the Low-Intensity Potential Threat phase. From 19 to 25 seconds, the administrator gestured to the child to pause in front of the unknown object while a repeated beeping sounded to increase a sense of urgency. This phase is meant to induce a High-Intensity Potential Threat response, such as startle or freezing behaviors, and is defined as the High-Intensity Potential Threat phase. From 25 to 30 seconds, the administrator removed the blanket revealing an empty box; and then debriefed the child, stating, “Oh, there’s nothing in there!”. This phase is meant to allow for Response Modulation such that the child can recover following disconfirmation of threat and is defined as the Response Modulation phase (Figure 1). Administration of the Approach Task is aided by a mobile application (ChAMP App) running on a smartphone that is worn on a belt around the child’s waist (42). The ChAMP app standardizes the start of the task, the timing of beeping associated with the task phases, and task end.

## Data analysis

### Wearable signal analysis

The BioStamp sensor recorded accelerometer and gyroscope data at a sampling rate of 250 Hz. A half-second bout where the child was standing upright and still was identified by applying a sliding window across the signals and selecting the interval with the lowest combined normalized variance of accelerometer and gyroscope signal magnitude, serving both for gyroscope bias removal and for anatomical calibration purposes. Mean angular velocity of the still bout was subtracted from the gyroscope signals throughout the approach task to remove gyroscope bias. The mean accelerometer measurement during the bout was leveraged to perform an anatomical calibration where a rotation matrix was created using Rodrigues’s rotation formula and applied to the sensor data so that measurements could be expressed in an anatomical reference frame with one axis aligned with the child’s cranial-caudal axis. Angular velocity in this cranial-caudal direction was extracted for further analysis. The Madgwick filter (43) was used to determine the orientation of the anatomical reference frame throughout the Approach Task by integrating the angular velocity while correcting drift with accelerometer measurements. Orientation at each timepoint during the task was computed as a quaternion and then converted to Euler angles (roll, pitch, yaw) for further analysis. The yaw Euler angle was baseline-corrected so that the first measurement at the beginning of the Low-Intensity Potential Threat phase was set to zero, capturing relative change in orientation from the beginning of this phase when the child is still oriented at the unknown threat.

We define Turning Speed and Turning Angle as the mean absolute value of the cranial-caudal angular velocity and yaw angle across the Approach Task, respectively. Turning Speed and Turning Angle are dynamically coupled signals, such that the integration of Turning Speed over time approximately yields Turning Angle. However, by considering both of these measures, we can capture the acute behavioral response of the child to the potential threat (Turning Speed: larger Turning Speed suggests a larger response), as well as how those responses aggregate over time into vigilance or avoidance behaviors (Turning Angle: smaller Turning Angle suggests more vigilance and larger Turning Angle means more avoidance).

## Statistical analyses

(1) Task Validity: Turning Speed was averaged during each task phase (Low-Intensity Potential Threat, High-Intensity Potential Threat, Response Modulation), for each child, to capture acute behavioral responses that we expected to differ based on threat intensity. Repeated-measures ANOVAs with post-hoc Bonferroni-corrected pairwise comparisons were used to determine whether subject behavioral responses differed across the task phases. Bonferroni correction was used to correct for multiple comparisons. (2) Clinical Validity: Turning Angle was averaged during the High-Intensity Potential Threat phase to capture the total threat response for each child. Linear regression was used to model this marker of total threat response using main effects of internalizing (CBCL internalizing T-score) and externalizing (CBCL externalizing T-score) symptoms and their interaction, adjusting for child age and sex.

To support interpretable visualization of these data, we developed groups based on symptom median splits. Children with CBCL T-scores above the medians are defined as High Externalizing (High Ext, CBCL externalizing T-score > 53) and High Internalizing (High Int, CBCL internalizing T-score > 54), while those with CBCL T-scores at or below the medians are defined as Low Externalizing (Low Ext) and Low Internalizing (Low Int) (Table 1).

**Table 1.** Subject demographics by symptom groups. Mean (Standard Deviation) of CBCL Internalizing and Externalizing T-scores by group. Int: Internalizing; Ext: Externalizing; CBCL = Child Behavior Checklist.

| Symptom Groups | N | CBCL T-scores |  |
| --- | --- | --- | --- |
|  |  | Internalizing<br>Mean (Std. Dev) | Externalizing<br>Mean (Std. Dev) |
| High Int & High Ext | 29 | 64 (6.2) | 64 (6.6) |
| High Int & Low Ext | 15 | 63 (7.8) | 49 (4.0) |
| Low Int & High Ext | 12 | 50 (2.7) | 60 (6.2) |
| Low Int & Low Ext | 35 | 44 (7.4) | 42 (6.6) |

## Results

### Acute behavioral responses confirm Approach Task validity

A repeated-measures ANOVA was used to compare behavioral responses across the three phases of the Approach Task to confirm that the task can serve as a behavioral probe of potential threat. The within-subject test revealed a significant main effect of task phase on mean Turning Speed (*F*(2, 86) = 45.2, *p* < .001, *η*^*2*^ = .342, Fig 2). Post-hoc Bonferroni-corrected pairwise comparisons indicated that behavioral responses (mean Turning Speed) were significantly larger during the Low-Intensity Potential Threat phase than both the High-Intensity Potential Threat (*MD* = 0.059, *SE* = 0.006, 95% *CI* [0.043, 0.075], *p* < .001) and Response Modulation (*MD* = 0.060, *SE* = 0.007, 95% *CI* [0.042, 0.077], *p* < .001) phases, with no difference between the High-Intensity Potential Threat and Response Modulation phases (Figure 2).

**Fig 2.**
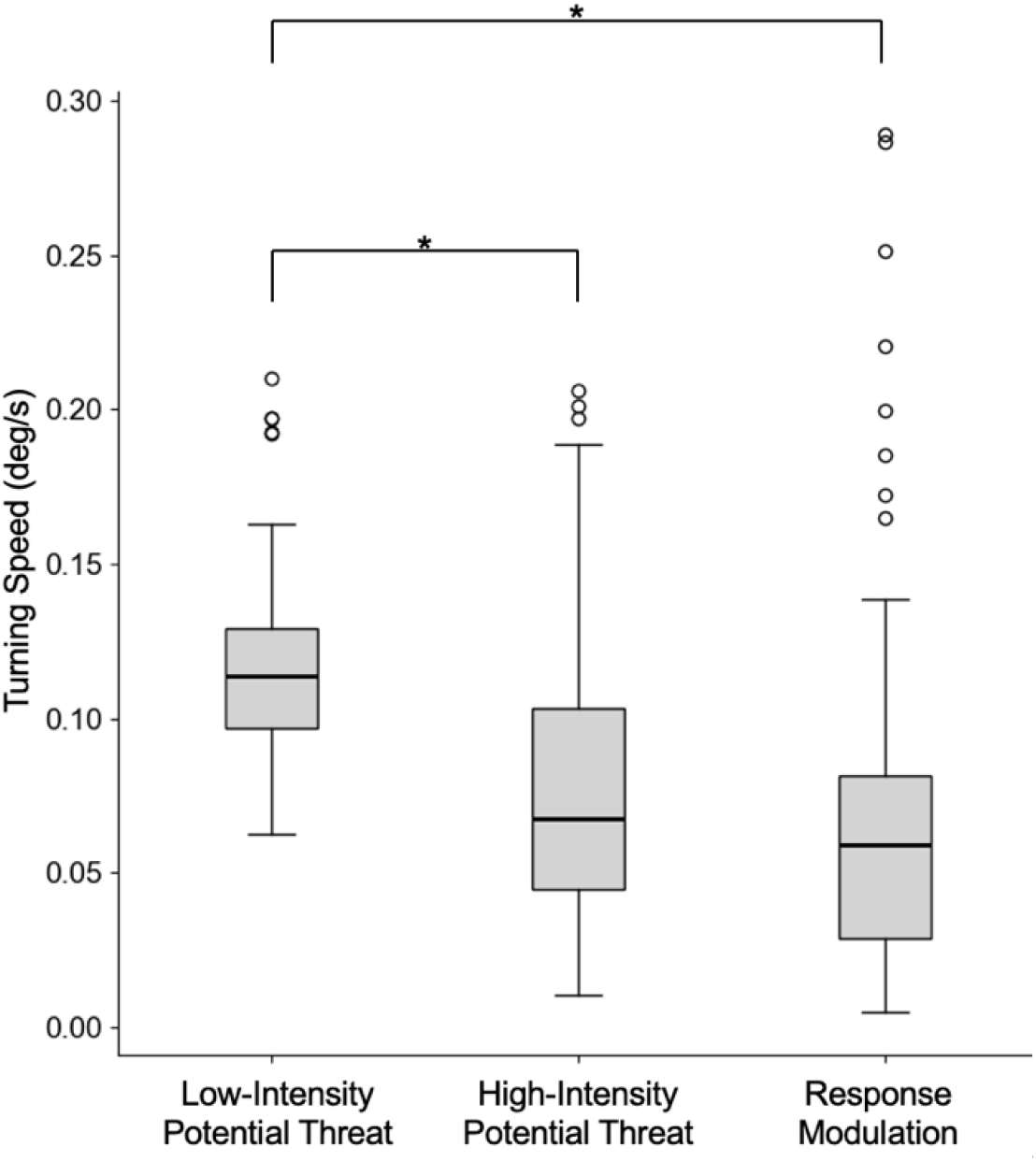
Boxplots of Turning Speed across all subjects during the three phases of the Approach Task: Low-Intensity Potential Threat, (5-19s), High-Intensity Potential Threat (19-25s), and Response Modulation (25-30s). Asterisks indicate statistically significant differences between phases as determined by post-hoc tests correcting for multiple comparisons (*p* < 0.05).

### Internalizing symptoms associated with threat response

Linear regression was used to explore associations between internalizing and externalizing symptoms and total threat response behavior during the Approach Task. The overall model (Table 2) was statistically significant (*F*(5,80)=2.39, *p*=.045), explaining 13% of the variance in mean Turning Angle during the High-Intensity Potential Threat phase (*R*^*2*^ = 0.13, Table 2). There was a significant main effect of CBCL internalizing T-score (*b*=−4.89, *SE*=2.12, *t*=−2.30, *p*=.024), such that higher internalizing symptoms were associated with a more vigilant threat response (smaller overall Turning Angle). Externalizing symptom severity, age, and sex were not associated with a threat response (*p*=.174, *p*=.531, and *p*=.986, respectively). However, there was also a significant interaction between the CBCL internalizing and externalizing T-scores (*b*=0.08, *SE*=0.04, *t*=2.03, *p*=.045), suggesting that the externalizing symptom severity moderates the effect of internalizing symptom severity on threat response behaviors.

**Table 2.** Associations of Turning Angle during the high-intensity potential threat phase of the Approach Task with child internalizing and externalizing symptom severity (CBCL T-scores).

| Independent Variable | $\beta$ (SE) | $t$ | $p$ |
| --- | --- | --- | --- |
| Intercept | 233.66 (113.80) | 2.05 | 0.043 |
| Internalizing Severity | -4.89 (2.12) | -2.30 | 0.024* |
| Externalizing Severity | -3.12 (2.28) | -1.37 | 0.174 |
| Internalizing x Externalizing Severity | 0.08 (0.04) | 2.03 | 0.045* |
| Age | 2.52 (4.00) | 0.63 | 0.531 |
| Sex | 0.17 (9.13) | 0.02 | 0.986 |

\* $p < 0.05$ .

To help visualize these findings, we present Turning Angle across the Approach Task by internalizing and externalizing severity groups in Figure 3. Specifically, children with high internalizing scores and low externalizing scores had smaller Turning Angles, whereas children with high internalizing and externalizing scores had larger Turning Angles.

**Fig 3.**
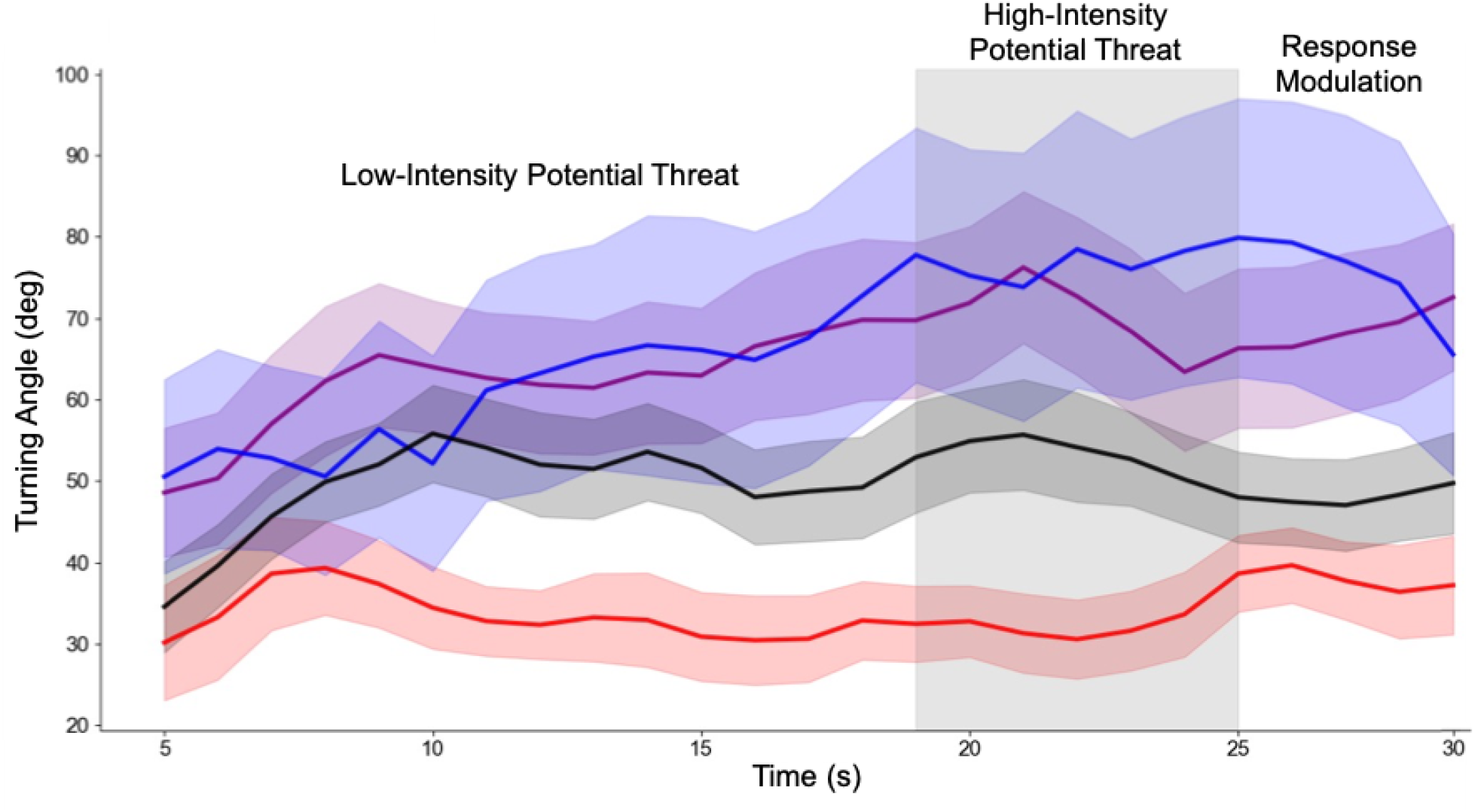
Turning Angle across the Approach Task for children with high internalizing and high externalizing (purple), high internalizing and low externalizing (red), low internalizing and high externalizing (blue), and low internalizing and low externalizing (black) symptom severity based on median splits from Table 1.

## Discussion

In this study, we examined Turning Angle and Turning Speed during a 30-second, potential threat paradigm (Approach Task) in relation to internalizing and externalizing symptom severity. Our aims were to (1) assess the task validity by examining changes in child movement in response to stimuli across the task and (2) assess clinical validity by examining associations between behavioral responses during the task and dimensional measures of internalizing and externalizing symptom severity and their interaction.

Phase-specific analyses revealed that child movement varied across phases of the Approach Task, consistent with our first hypothesis. Turning Speed was faster during the Low-Intensity Potential Threat phase, which then decreased during the High-Intensity Potential Threat phase, remaining stable during the Response Modulation phase. The movement change between Low and High-Intensity phases may be consistent with literature showing distinct brain activation between potential and more acute threat (44). Alternatively, this pattern may reflect preliminarily exploratory behavior when first encountering an unfamiliar, potentially threatening environment, followed by more cautious or inhibited movements as the threat became more immediate. These results align with previous research showing that children exhibit heightened vigilance and exploratory movement during ambiguous or low-threat conditions, then transition to freezing or reduced motion as the perceived threat increases (18,45). Importantly, these dynamic changes were captured by a single wearable sensor on the child’s lower back, supporting the construct validity of the Approach Task and demonstrating that objective, sensor-based movement features can provide insight into young children’s threat response according to threat intensity.

Children with higher internalizing symptom severity (and low externalizing symptom severity) exhibited smaller Turning Angles suggesting a threat response of orienting toward the stimulus. This finding is consistent with our second hypothesis and previous literature (e.g., anxiety, social withdrawal) (23,36,46,47), reflecting sustained vigilance for potential threat. Inconsistent with prior literature (19,32), we found that externalizing symptom severity alone was not associated with Turning Angle, when it is typically associated with orientation away from the stimulus. However, we found that children with comorbidity (high internalizing and high externalizing symptoms) displayed greater Turning Angles, suggesting orientation away from threatening stimuli (32). In contrast to one prior study observing that children with high comorbid anxiety and ADHD symptoms exhibited an anxiety-only-like threat response, we found that the opposite threat response prevailed (Fig. 3). Here, larger Turning Angles could reflect avoidance behaviors. The vigilance-avoidance model of attention (20,21,26,48) posits that initial vigilance to detect threat can lead to subsequent avoidance strategies. We could speculate that children with high comorbid symptoms have more impaired functioning and could have had to develop and use avoidance strategies earlier in life than children with high internalizing symptoms only. Alternatively, children with high comorbid symptoms could have larger Turning Angles due to symptoms of hyperactivity (32), emphasizing the need to examine interactions between negative valence (i.e., threat sensitivity) and response modulation systems (i.e., behavioral control and inhibition) (41). Comorbid externalizing symptoms appear to alter behavioral expression of anxiety-related processes, highlighting the importance of considering comorbid symptom profiles in future studies.

Our findings extend prior work by validating a brief assessment of potential threat for young children. We consider how comorbid symptoms modulate behavioral responses (1), enhancing ecological validity by including children who span symptom severity (1,2,34,41) and demonstrate that less than 30 seconds of movement data measured by a wearable sensor may yield behavioral biomarkers for childhood mental health risk that could complement traditional assessments relying on parent reports.

Limitations of this study include the relatively small and demographically homogenous sample, which limits generalizability. Furthermore, we consider movement metrics as averages over relatively long task phases relative to the frequency of movements within those phases, which may obscure meaningful shorter-term movement patterns in Turning Speed. Similarly, relying on task initialization to define baseline child orientation could add additional variability to the measurements of Turning Angle. Turning speed and angle are only two possible features that could be associated with internalizing and externalizing symptoms. Future research should examine turning behaviors when presented with a potential threat in a larger and more diverse sample of children and leveraging alternative processing methodologies to further explore potential correlates of internalizing and externalizing symptoms. Finally, combining wearable-based movement measures with other physiological data (e.g., heart rate, electrodermal activity, etc.) may provide a more comprehensive understanding of potential threat responses and reveal stronger associations with symptom profiles.

## Data Availability

The datasets presented in this article are not readily available because they contain sensitive personal information derived from clinical interviews and wearable sensors. Because of institutional and legal restrictions related to privacy protection, the data cannot be shared publicly or upon request. Requests to access the datasets should be directed to

## Conclusion

Our findings identify key movement features derived from a single wearable sensor that reflect threat behavioral responses in young children during a 30-second Potential Threat paradigm. Threat responses were associated with internalizing symptom severity and moderated by comorbid externalizing symptom severity. This approach highlights the promise of scalable, objective tools for assessing threat response behaviors in early childhood and supports the application of dimensional frameworks to understand the interaction of comorbid symptoms on behavioral responses.

## Acknowledgements

The authors thank the children and families who participated in the study. Fig. 1 created with BioRender.com.

